# Statistical Analysis Plan for the Optimal Post rTpa-Iv Monitoring in Ischemic Stroke Trial (OPTIMIST)

**DOI:** 10.1101/2024.12.17.24319094

**Authors:** Laurent Billot, Severine Bompoint, Menglu Ouyang, Lizheng Xu, Xia Wang, Alejandra Malavera, Brenda Johnson, Debbie Summers, Pooja Khatri, Paula Muñoz-Venturelli, Diana Day, Yi Sui, Lili Song, Wan Asyraf Wan Zaidi, Nguyen Huy Thang, Candice Delcourt, Thompson Robinson, Stephen Jan, Richard I Lindley, Victor C Urrutia, Craig S Anderson

## Abstract

1

The OPTIMIST trial aims to determine whether low-intensity monitoring is at least as effective (“non-inferior”) to standard monitoring, on the functional recovery of patients who have received recombinant tissue plasminogen activator or equivalent lytic reperfusion treatment for acute ischaemic stroke. It is designed as an international, multicenter, stepped-wedge (4 periods/3 steps) cluster randomized trial. This statistical analysis plan pre-specifies the method of analysis for every outcome and key variable collected in the trial.

The primary outcome is an “unfavourable outcome” at Day 90, defined as a score of 2 to 6 on the modified Rankin scale. The primary analysis will consist in a log-binomial regression adjusted for the effect of time and for clustering by site using a random effect. The non-inferiority margin was pre-specified as a relative risk of 1.15 for a bad outcome; thus, non-inferiority will be declared if the upper bound of the 95% confidence interval around the relative risk is lower than 1.15.

The primary analysis will adjust for calendar time (6-month intervals) and will be based on imputed data. The analysis plan also includes planned sensitivity analyses including covariate adjustments and subgroup analyses.

**Administrative information:** *Study identifiers:*

- Protocol Number: GI-AU-NMH-2019-CA Version: 3.0, Date: 3 March 2021
- ClinicalTrials.gov register Identifier: NCT03734640
- Australian New Zealand Clinical Trial Registry: ACTRN 12619001556134

*Revision history:* 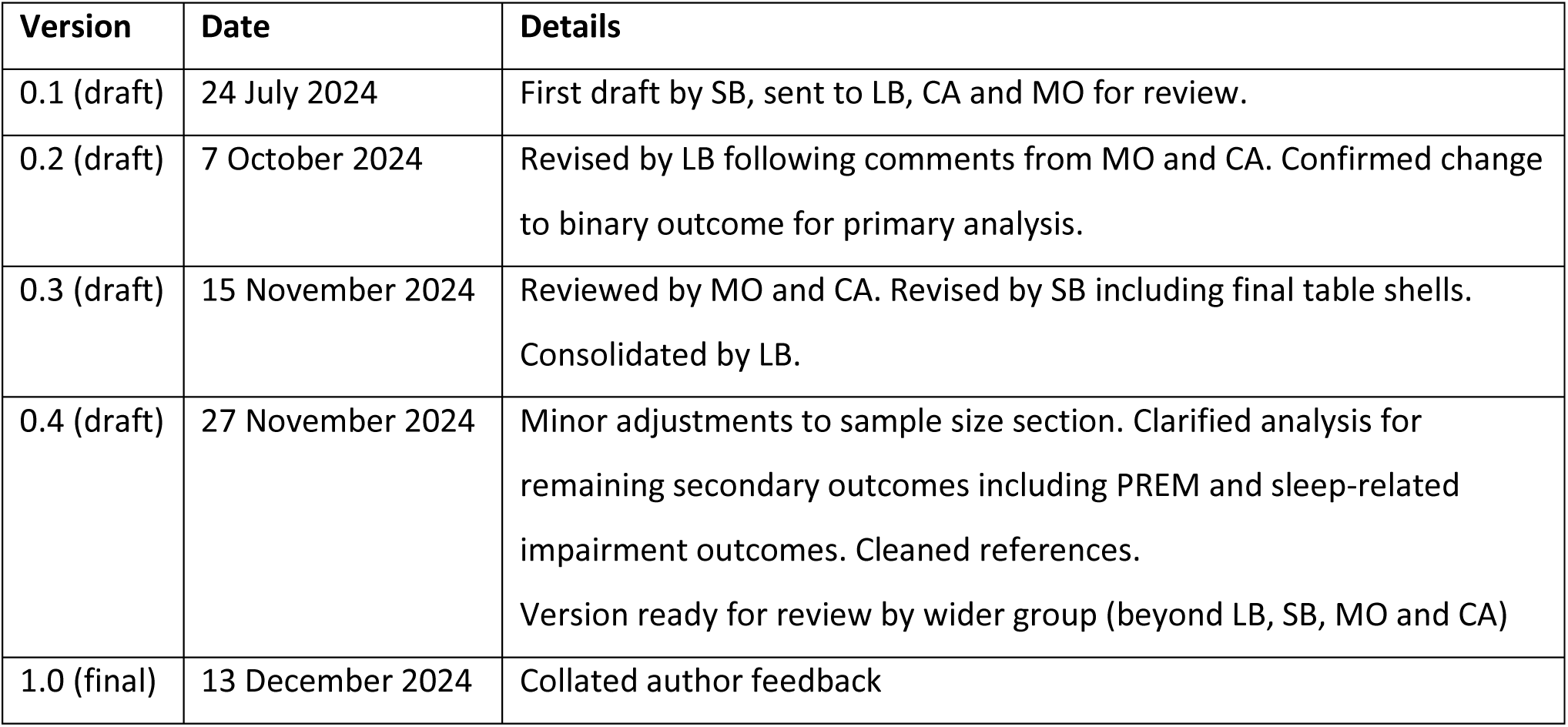

*Contributors to the statistical analysis plan:* LB – Main author; developed the initial draft and prepared subsequent versions. SB - Prepared initial draft and reviewed subsequent versions. MO, CA - Reviewed every draft and approved final version. LX - Prepared economic analysis section and approved final version. SJ - Comments on economic analysis plan; approved final draft. XW, AM, BJ, DS, PK, PM-V, DD, YS, LS, WAWZ, NHT, CD, TR, RIL - Approved final draft.

## 3 Introduction

### 3.1 Study synopsis

The OPTIMISTmain clinical trial is an investigator-initiated and conducted, regionally coordinated, international, pragmatic, multicentre, prospective, stepped-wedge, cluster randomised, blinded outcome assessed study being undertaken through a global network of investigators.

The key objective is to determine whether low-intensity monitoring is at least as effective (‘non-inferior’) to standard monitoring, on the functional recovery of patients who have received recombinant tissue plasminogen activator (rtPA) or equivalent lytic reperfusion treatment for acute ischaemic stroke (AIS). Another aim is to establish whether such low-intensity monitoring is safe, and provides economic and resource benefits, relative to standard monitoring. The full protocol was published in 2023.[1]

Patient enrolment commenced between 28 April 2021 and 30 September 2024. As of 30 September 2024, there were 4834 patients enrolled at 118 hospital sites in Australia, China, Chile, Malaysia, Mexico, Vietnam, United Kingdom (UK), and the United States of America (US). Patient follow-up will be completed by 30 December 2024, and the study closed by 31 January 2025. The main results will be published in May 2025.

### 3.2 Study population

#### 3.2.1 Site eligibility criteria

Hospital sites were eligible if they had an established acute stroke care program / pathway, and an acute stroke unit (or a geographically-defined organised area) for the management of patients with acute stroke. Sites must have agreed to implement the intervention alongside an established stroke care program and to have the capacity to enter data onto a secure web-based server. It is imperative that the hospital investigators complied with the protocol and Good Clinical Practice (GCP) requirements, and to have had the capacity to appoint an appropriately trained independent person to undertake 90-day outcome assessments in a blinded manner, where possible.

#### 3.2.2 Patient eligibility criteria

- **Inclusion criteria**

`o` Adults (age ≥18 years; NB. the age for adults may vary in some countries)
`o` A clinical diagnosis of AIS
`o` Given intravenous (IV) bolus/infusion of rtPA or equivalent thrombolytic agent
`o` Vital signs are stable with a mild-moderate level of neurological deficit (e.g. National Institutes of Health Stroke Scale [NIHSS] score <10) within 2 hours post-rtPA
- **Exclusion criteria**

`o` Definite contraindication for low-intensity monitoring, in the opinion of the treating clinician (i.e. requiring intensive care unit [ICU] admission and/or prolonged intensive monitoring for a severe neurological deficit, evidence of early neurological deterioration or perceived potential for further neurological deterioration, requiring airway support, or in need of specific intravenous therapy that cannot be given on a general ward);
`o` Immediate transfer for medical treatment (e.g. for haemodialysis; co-morbid conditions such as renal failure; palliative care) or surgery (e.g. carotid endarterectomy) where adherence to low-intensity monitoring is not possible.

In each case, the decision about the patient’s eligibility was based on the attending clinician investigator’s interpretation of the above eligibility criteria.

### 3.3 Interventions

#### 3.3.1 Intervention arm

The intervention was to be an allocated monitoring strategy for all eligible post-rtPA AIS patients during the study period, to have been started immediately following thrombolysis, which are: vital signs (HR, BP) and neurological assessment (Glasgow coma scale [GCS] and/or NIHSS) 15 mins x 2 hours at the beginning of rtPA infusion, then 2 hourly x 8 hours, and 4 hourly x 14 hours.

#### 3.3.2 Usual Care arm

As soon as a site was activated, patients in the Usual Care group received standard of care which consists of vital signs (HR, BP) and neurological assessment (GCS and/or NIHSS) 15mins x 2 hours at the beginning of rtPA infusion, and then 30mins x 6 hours, and hourly x 16 hours - as per recommended guidelines

#### 3.3.3 Background standard of care

All background care of AIS patient care post-rtPA should continue as per usual care at that site. The focus of the experimental intervention is on low-intensity monitoring, but sites were encouraged to consider undertaking the intervention on a stroke unit ward rather than in ICU, provided that was permitted within the organisational structure of the hospital.

### 3.4 Outcomes

The study outcomes are:

#### 3.4.1 Primary outcome

- Unfavourable (disabled or dead) functional outcome, defined as a score of 2 to 6 on the modified Rankin scale (mRS) score at Day 90.

#### 3.4.2 Secondary outcomes

- Functional outcome defined as an ordinal assessment across all 7 levels of the mRS at Day 90
- Investigator-reported symptomatic intracerebral haemorrhage (sICH) according to Heidelberg criteria.
- Requested time to CT scan for presumed sICH from last physiological monitoring
- Death or dependency measured by a shift in NIHSS scores at 7 days or before discharge if earlier
- Death within 90 days
- Bad functional outcome (mRS 3-6 vs 0-2) at 90 days
- Length of hospital stay at 7 days or before discharge if earlier
- Health-related quality of life (HRQoL) on the European Quality of Life Scale 5 Dimension, 5L Version (EQ-5D-5L) and EQ-VAS at 90 days
- Patient-reported experience measurement (PREM) at 7 days
- Sleep-related impairment at 7 days

#### 3.4.3 Safety outcomes

- Serious adverse events (SAEs) during follow-up

### 3.5 Randomisation and blinding

The unit of randomisation was the site, randomly assigned by an unblinded statistician not otherwise involved in the study, into one of the three groups using a pre-specified randomisation stratified according to country and size of the hospital (by patient number of annual admission for thrombolysis treatment <=60 or >60). All AIS patients eligible for thrombolysis were prospectively and consecutively enrolled.

### 3.6 Statistical hypotheses

The primary statistical hypotheses are as follows:

- **Null hypothesis**: low-intensity monitoring is not as effective as standard monitoring, on the functional outcome of patients who have received thrombolysis treatment for AIS (RR of a bad outcome of 1.15 or more).
- **Alternative hypothesis (1-sided)**: Low-intensity monitoring strategy vs standard monitoring strategy will be as effective (‘non-inferior’), on the functional outcome of patients who have received thrombolysis treatment for AIS (RR of a unfavorable outcome less than 1.15).

### 3.7 Sample size

In a key individual patient meta-analysis from the Stroke Thrombolysis Trialists Collaborative Group, rtPA increased the odds of an excellent clinical outcome (defined by mRS scores 0–1) compared to control (36.7% versus 29.1%) in 3491 patients who met the age-revised European Union label.[2] This corresponds to an odds ratio 1.43 (95% confidence interval [CI] 1.23-1.65) or a RR of 1.26 (95%CI 1.15-1.40). We chose the lower 95% CI boundary (i.e. 1.15) of the anticipated treatment of rtPA to establish the non-inferiority margin to correspond to loss of efficacy of rtPA from low-intensity monitoring.

Assuming patients have a 50% chance of experiencing an unfavourable outcome (mRS 2-6) in both arms (i.e. an actual RR of 1.0) as observed in the Enhanced Control of Hypertension and Thrombolysis Stroke Study (ENCHANTED),[3] an individual randomised clinical trial of 2,164 participants would provide 90% power (one-sided α = 0.025) to reject a non-inferiority margin of 1.15 for the relative risk of an unfavourable outcome. A non-inferiority margin of 1.15 on the relative scale represents a 7.5% absolute increase in the proportion of patients with a bad outcome (from 50% to 57.5%).

Using a step-wedged design of three groups and three steps (four periods), with each hospital recruiting an average of 15 patients per period, a total of 108 hospitals are needed with 3 groups of 36 hospitals. Assuming there is 10% of participants with missing primary endpoint data, we aimed to recruit 16 participants per hospital per period, for a total of 6,912 participants. This calculation assumes: (i) there is a similar treatment effect from the low-intensity monitoring strategy as the low-dose rtPA-IV; and (ii) an interclass correlation coefficient (ICC) of 0.044 between sites which is similar to that found in the international Head Position in Acute Stroke Trial (HeadPoST).[4]

## 4 Statistical analysis

### 4.1 Statistical principles

#### 4.1.1 Level of statistical significance

Two formal interim analyses were planned with the use of the Haybittle-Peto stopping rule (3 standard-deviations) for efficacy,[5] but these were not undertaken due to the imbalance in participant numbers between randomized groups inherent to the stepped-wedge design (the control Usual Care arm was completed before all the sites had crossed over to the intervention). Moreover, China contributed a large proportion of participants but only in the final year of the study. Given the absence of overall (blinded) safety concern, no formal interim analysis was conducted. Thus, the significance threshold will remain at 5% for the final analysis.

Final analyses of the primary outcome, including sensitivity analyses, will all be conducted using a two-sided significance level of 5%.

This is a non-inferiority study which assumes that low-intensity monitoring has a similar effect to standard monitoring. We do not expect significant differences between the two arms across secondary outcomes. The analysis of secondary outcomes will therefore focus on point and interval estimation rather than significance testing. For these reasons, we will not adjust for multiplicity and will not report p-values for the secondary and other outcomes.

#### 4.1.2 Statistical software

Analyses will be conducted primarily using SAS (version 9.3 or above) or R (version R-4.3.1 or above).

### 4.2 Analysis populations

Due to the stepped-wedge design, the group allocation for a patient was determined by the site and by the period during which they participated, regardless of treatment adherence.

The ***intention-to-treat (ITT) analysis set*** will be used to assess both effectiveness and safety. The flow of patients through the study will be displayed in a CONSORT diagram (Figure 1). Details will be obtained as to the type of assessment performed (phone, face-to-face, etc.) and analyses will be undertaken according to recent reporting guidelines for stepped-wedge cluster randomised trials.[6]

The ***per-protocol (PP) analysis set*** will be defined as patients in whom the allocated monitoring protocol was followed unless the reason for change was a medical event. Patients included in the PP populations will be those who responded “Yes” to “Was study allocated monitoring post-thrombolysis protocol followed?” (Question 5 in Figure 1) or those who ticked “Medical event” or “Neurological deterioration” as the reason for interruption of the monitoring care” (Question 19 in Figure 1). The PP set will be used to rerun analyses of the primary outcome, including sensitivity analyses (see Section 4.8), as well as the analysis of SAEs (Section 4.9.8).

**Figure 1.**
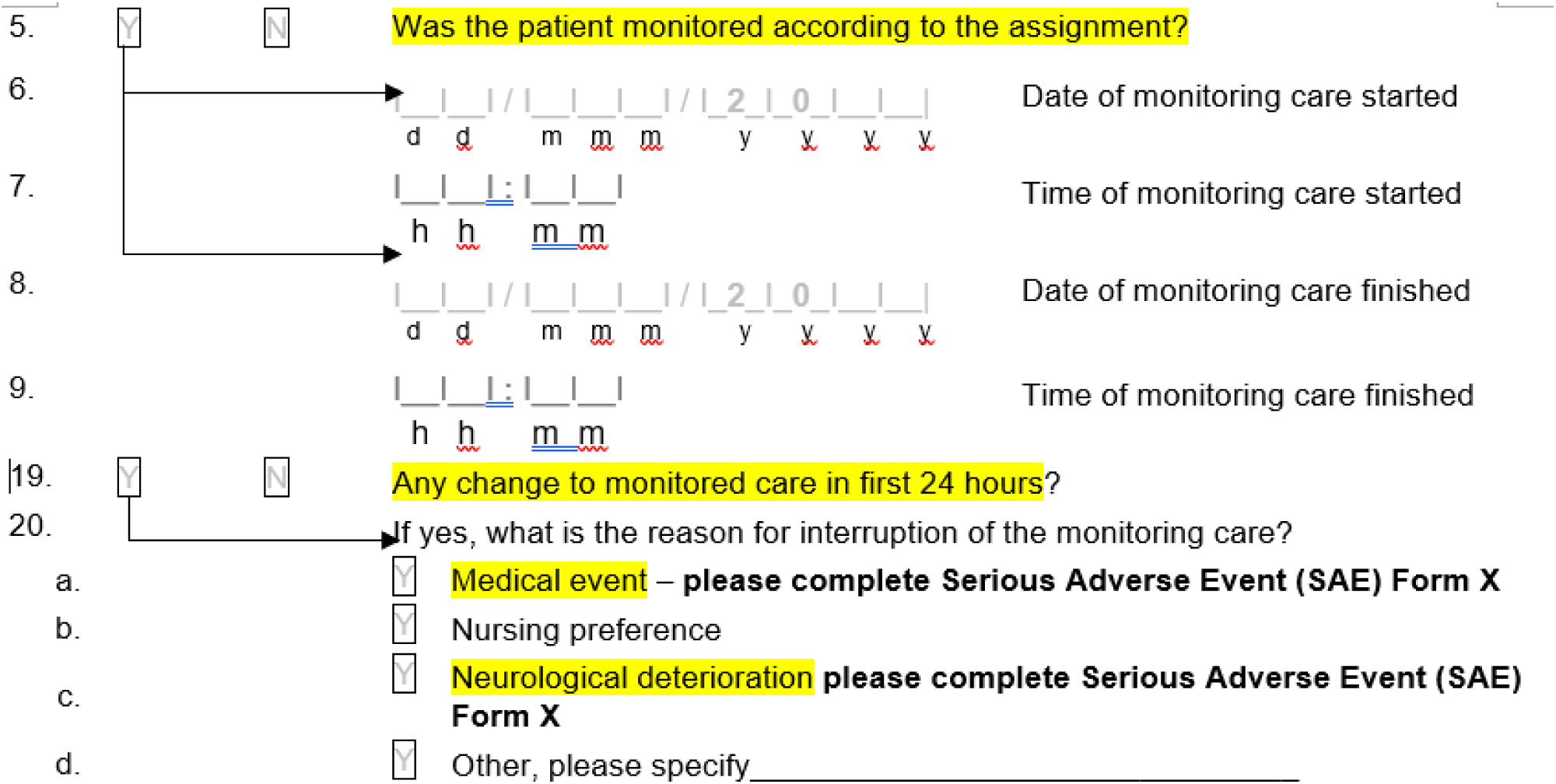
Extract from case report form C.

All analyses will be run in the ITT set. Only the analyses of the primary outcome (Section 4.8.1), including sensitivity analyses (Section 4.8.3), will be repeated in the PP analysis set. Subgroup analyses will not be repeated in the PP analysis set. Further PP analyses and other analyses adjusting for compliance will be conducted as part of secondary publications.

### 4.3 Impact of COVID

The COVID pandemic, which emerged during conduct of the trial, had a significant impact on recruitment in many countries. We will perform adjusted analyses by including time and key baseline characteristics (age, sex and NIHSS) to account for the possible impact of COVID on patient profiles and other secular change in background care.

### 4.4 Changes from the protocol

This SAP is based upon Version 3.0 (3 March 2021) of the protocol. However, following a blind review of the grouped data and a review of recent literature, we will implement the following changes:

1. **Primary analysis**: in the protocol, we indicated the primary analysis will consist of an ordinal analysis of mRS at 90 days using an ordinal logistic regression to produce a common odds ratio (cOR). Recent trials,[7,8] however, model the probability of an excellent outcome (mRS 0-1) using a binary logistic regression. Given the skewed distribution of mRS scores at 90 days (approximately two-thirds of patients have a score of 0-1 at the time of writing this SAP) and the challenges associated with using OR in non-inferiority analyses, we have changed the primary outcome to a binary outcome (mRS 2-6 vs 0-1) and the primary analysis will be with a log binomial regression. Adjusted absolute differences and 95% confidence intervals (CI) will be obtained from the regression model and used to test the non-inferiority hypothesis. The non-inferiority margin will be based on a margin of 1.15 for the relative risk of an unfavourable outcome (mRS 2-6) as per the original protocol. The sample size calculation has been adjusted slightly to reflect the use of RR as the non-inferiority criteria.
2. ^1^**Time adjustment:** In the published protocol, we stated that a blind review would inform the choice of approach for the modelling of time trends, using either study period or calendar time. A blind review of start and stop times by site and period revealed substantial variation in the timing and duration of periods across sites. We have therefore decided to perform the primary analysis by adjusting for calendar time instead of study period.
3. **Missing data imputation:** The published protocol mentions controlled multiple imputations as a sensitivity analysis. Given that we expect between 10% and 15% of missing data, we have decided that the primary analysis would be based on imputed data using multiple imputations.

All these changes are made in a blinded manner and independent of any trial result.

### 4.5 Baseline analyses

#### 4.5.1 Cluster characteristics

Description of the cluster characteristics (e.g. location, size) will be presented by treatment group. Discrete variables will be summarised by frequencies and percentages. Percentages will be calculated according to the number of clusters with available data. Continuous variables will be summarised by using mean and standard deviation (SD), and median and interquartile range (Q1-Q3).

#### 4.5.2 Patient characteristics

Description of the baseline characteristics will be presented by treatment group and by study period. Discrete variables will be summarised by frequencies and percentages. Percentages will be calculated according to the number of patients in whom data are available. Continuous variables will be summarised by using mean and SD, and median and interquartile range (Q1-Q3). No adjustment for clustering will be applied when summarising baseline characteristics.

Baseline measures will include all socio-demographic, clinical and medical information collected at baseline.

### 4.6 Assessments and interventions conducted between Day 1 and Day 7

All assessments performed and interventions received between Day 1 and Day 7 will be described by intervention group. No formal statistical tests are planned for these variables.

Protocol deviations will be categorised and reported as the number and proportion of subjects experiencing a deviation. A listing of all protocol deviations will be provided.

### 4.7 Vital signs

BP and HR values collected at randomisation and at Day 7 will be summarised using box plots and by reported as mean and standard deviation (SD), and median and interquartile range (Q1-Q3) by visit.

### 4.8 Analysis of the primary outcome

The primary outcome is an “unfavourable outcome” at Day 90, defined as a score of 2 to 6 on the mRS. It will be analysed using log-binomial regression adjusted for the effect of time and for clustering by site. The effect of the intervention will be estimated as a relative risk (RR) and its 95% CI. The non-inferiority margin was pre-specified as a relative risk of 1.15 for a bad outcome; thus, non-inferiority will be declared if the upper bound of the 95%CI around the RR is lower than 1.15. The primary analysis will adjust for calendar time (6-month intervals) and will be based on imputed data.

#### 4.8.1 Main analysis of the primary outcome

The primary analysis will be conducted following the approach developed by Hussey and Hughes for the analysis of stepped-wedge trials.[9] Given that the non-inferiority margin is expressed in terms of RR, we will be using log-binomial regression to derive an adjusted risk ratio instead of an OR. This will be done using a generalised linear mixed model assuming a binomial distribution and a log link with a fixed categorical effect of time and a fixed effect indicating the group assignment of each participant (at their time of inclusion / randomisation). To account for within-hospital clustering, the model will include a random hospital effect.

Given substantial variations in period start times and durations across hospitals, the effect of time will be modelled using calendar time instead of study period. Time will be defined as seven 6-month intervals starting from 28 April 2021 (first patient recruited) and finishing in 30 September 2024 (last patient recruited).

The effect of the intervention will be derived as the RR of an unfavourable outcome (mRS 2-6) and its 95%CI using the standard arm as the reference (i.e., where a RR greater than unity corresponds to an increase in bad outcomes in the low-intensity arm compared to the standard arm). The low-intensity intervention will be declared non inferior to standard care if the upper bound of the 95% CI around the RR is lower than 1.15. If non-inferiority is declared, we will subsequently test for superiority (upper bound of the 95%CI lower than 1). P values associated with both the non-inferiority and superiority hypotheses will be derived from the analysis model.

Randomisation was stratified by country and size of hospital. Given the large number of strata, and that the primary analysis is already adjusted by site, the main analysis model will not further adjust for country or size of site. Further adjustments will, however, be performed as sensitivity analyses (see Section 4.8.3.3).

Given that we expect more than 10% of patients with missing primary outcome data, the primary analysis will be performed using multiple imputations as described in Section **Error! Reference source not found.**.

In case of convergence issues with the log-binomial regression, we will use robust Poisson regression (i.e. assuming a Poisson distribution) instead of a binomial one and using an empirical “sandwich” estimator to estimate covariance parameters in order to correct for the misspecification of the distribution.[10] Furthermore, in case of convergence issues with the random effect, the within-cluster correlation will be modelled using a repeated effect instead of a random effect (i.e., by directly modelling correlations between errors for participants from the same cluster assuming an exchangeable correlation structure).

Except for subgroup analyses, analyses of the primary outcome, including sensitivity analyses, will be run in both the ITT and PP populations (see Section 4.2).

**Table 1.**
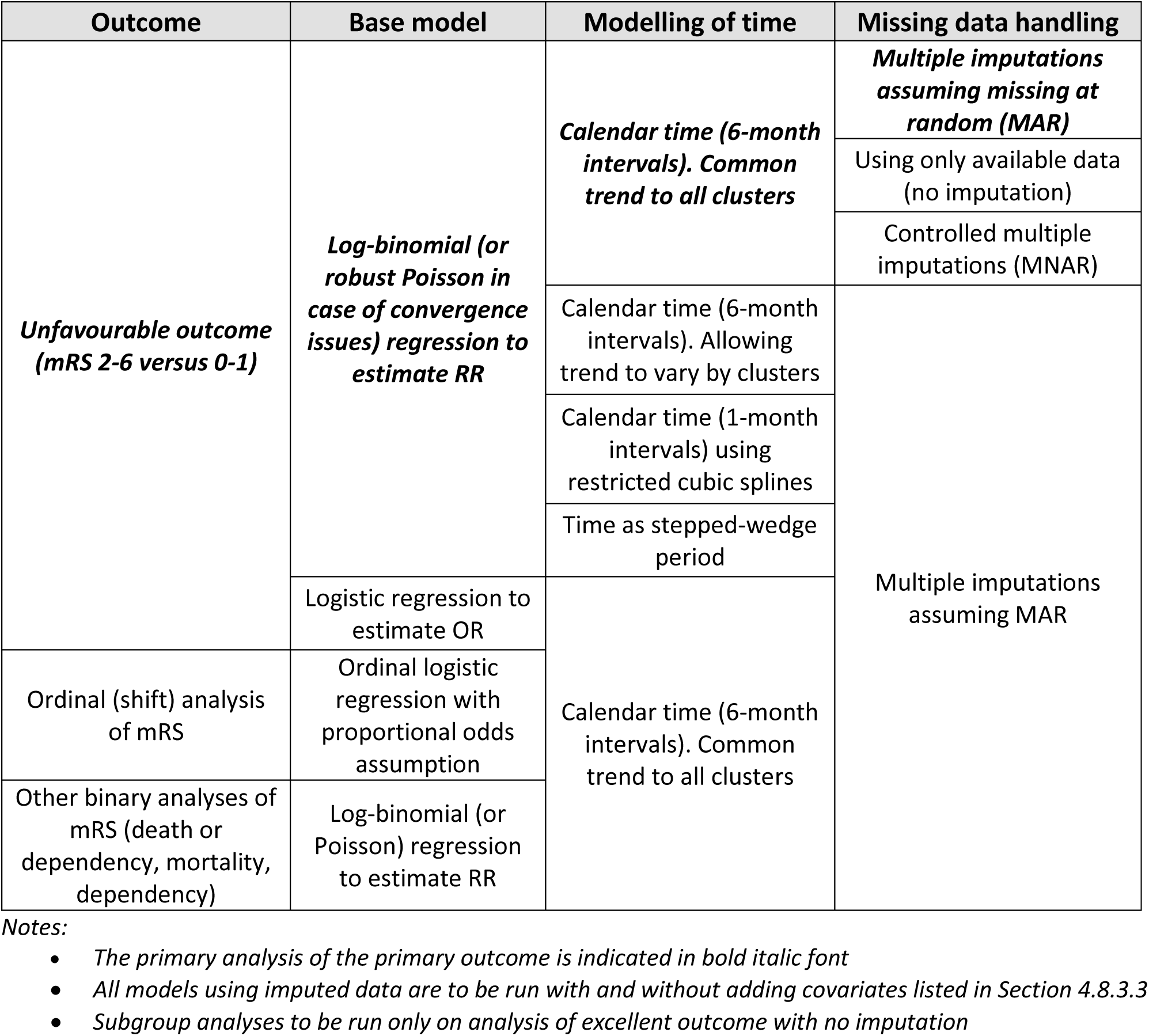
Summary of analyses of mRS at 90 days.

#### 4.8.2 Missing data handling

Following a blind review of the data, we estimate that between 10% and 15% of mRS at 90 days will be missing. The primary analysis will therefore be based on multiple imputations assuming that data are missing at random (MAR). Sensitivity analyses4.3 using controlled multiple imputations are described in Section 4.8.3.4.

We will use fully conditional specification (FCS) [11] and will include the following variables: mRS at Day 90, the NIHSS and mRS scores at 7 days (or hospital discharge, if sooner), a variable indicating the cluster (hospital), a variable indicating the calendar time period, a variable indicating the intervention, and all key socio-demographic, clinical, and medical baseline variables, as reported in Table , Table and Table . The mRS at Day 90 and NIHSS at 7 days will be imputed using an ordinal logistic model. Other variables will be imputed using either linear regression (for continuous / ordinal variables) or a discriminant function method (for nominal variables). One hundred sets of imputed data will be created and analysed using the model described in Section 4.8.1. Estimates of the treatment effect and its standard errors will be combined to obtain a pooled common RR and 95% CI.

For reference, and to assess consistency, a model will also be run without any missing data imputation (i.e. using all data available). This non-imputed model will be used as a basis for subgroup analyses (see Section 4.8.4).

#### 4.8.3 Sensitivity analyses

##### 4.8.3.1 Logistic regression

The main analysis described in Section 4.8.1 will be repeated using a logistic model instead of a log-binomial (or robust Poisson) model. This will be done by assuming a binomial distribution and a logit link and without the need for robust/empirical standard errors. The rest of the model including covariates, adjustment for clustering by hospital and the modelling of time will remain the same. This analysis will be done on the same imputed datasets. The effect of the intervention will be estimated as the odds ratio of an unfavourable outcome and its 95%CI. The purpose of this model is to check that the results obtained via logistic regression are consistent with the ones obtained from the robust Poisson model; however, this model will not be used to formally test the non-inferiority hypothesis.

##### 4.8.3.2 Allowing time trends to vary by cluster

The base model developed by Hussey and Hughes makes the implicit assumption that the effect of time is common across all clusters. Given the large number of clusters and the fact that sites entered the study at different times, an additional model will be run to allow the effect of time to vary randomly across sites, as described by Hemming et al.[12] This will be done by adding a random interaction between period and cluster.

The addition of a random period-by-cluster interaction will allow the intra-cluster correlation to vary depending on whether subjects from the same cluster are from the same period or from a different period; thus, modelling a two-period decay correlation structure instead of an exchangeable one. This model will otherwise be the same as the main model described in Section 4.8.1.

##### 4.8.3.3 Adjusted analyses

The models described in Sections 4.8.1 and 4.8.3.1 will be re-run after adding the following individual baseline covariates as fixed effects:

- Country (grouped as Australia, China, Malaysia + Vietnam, UK, Chile + Mexico, and US)
- Baseline mRS (categorical variable)
- Age (continuous)
- Sex (male vs female)
- Baseline NIHSS score (continuous)

In case of convergence issues with the fully adjusted model, we will either remove or collapse the categorical covariates starting with country.

##### 4.8.3.4 Controlled multiple imputations

To assess the robustness of the results under various assumptions about the missing data, we will perform controlled multiple imputations following the approach by Cro et al.[13]

Using the same “base” 100 sets of imputed data constructed for the main analysis (Section 4.8.2), we will assume different mRS levels for subjects who had missing mRS data at Day 90 and had their mRS value imputed. We will first assume that those with a missing mRS were more likely to have a poorer outcome than those with a non-missing mRS, and will add ‘1’ to their imputed mRS score (up to a maximum score of 6). We will then analyse the 100 modified-imputed datasets and combine the results using the same analysis strategy as for the base set of imputed data. As an additional sensitivity analysis, we will impute all missing mRS score at 90 days with a score of 6, thus assuming all subjects with missing data have died. While this assumption is unlikely to hold in most cases, it is plausible that some subjects may have become uncontactable due to death after hospital discharge. Depending on the results, additional scenarios might be explored to further test the robustness of the results. Controlled multiple imputations will only be applied to the primary outcome and only to the main analysis model described in Section 4.8.1. (i.e. Log-binomial (or Robust Poisson) regression with time modelled as calendar time in 6-month intervals and a common trend across all clusters).

##### 4.8.3.5 Alternative methods for modelling the effect of time

The primary model will model time as calendar time in 6-month periods. To assess the robustness of results to the way the effect of time is modelled, we will consider two alternative approaches:

1. Modelling calendar time using restricted cubic splines. Time will be categorised in 1-month intervals and modelled as a continuous covariate.
2. Modelling time as the stepped-wedge period as implemented at each site. Time will be categorised into 4 study periods.

These two approaches will only be applied to the main outcome analysis described in Section 4.8.1. No further covariate adjustments, controlled imputations or subgroup analyses, will be applied.

#### 4.8.4 Subgroup analyses

The following subgroup analyses are planned for the primary outcome, regardless of the statistical significance for the primary analysis:

- Age: <60, 60 to <70, 70 to <80, and 80+
- Sex: male vs. female
- Country: China vs. Malaysia/Vietnam vs. Chile/Mexico vs. Australia vs. UK vs USA
- NIHSS on arrival: <5 vs. ≥5
- Ethnicity: White/Caucasian vs. Asian vs. Hispanic vs. Black vs. all others
- AIS pathological subtypes (small vessel ‘lacunar’ vs large vessel vs cardio-embolic vs other cause)
- Location to receive post-thrombolysis care: acute stroke unit vs. intensive care unit vs. intermediate/step down/high dependency care unit vs. other

Each subgroup analysis will be performed by adding the subgroup variable as well as its interaction with the intervention as fixed effects to the Poisson regression model used for the primary analysis (see Section 4.8.1). Within each subgroup, summary measures will include raw counts and percentages within each treatment arm, as well as the RR for treatment effect with 95% CI. The results will be displayed on a forest plot including the P value for heterogeneity corresponding to the interaction term between the intervention and the subgroup variable. Given the main purpose of these subgroup analyses is exploratory and to assess the consistency of results, they will only be performed on the non-imputed data (i.e. using all data available). For subgroup analyses by country and ethnicity, categories might need be collapsed in case of estimation issues.

### 4.9 Analysis of secondary outcomes

#### 4.9.1 Ordinal analysis of mRS at Day 90

The method to analyse the mRS as an ordinal outcome will be similar to the binary analysis of mRS described in Section 4.8.1. However, instead of using Poisson regression, we will use an ordinal logistic regression assuming proportional odds and will not use robust/empirical standard errors. The effect of the intervention will be presented as the OR of a worse outcome and its 95% CI.

In case of violation of the proportional-odds assumption, we will proceed with the analysis and interpret the intervention OR as an average effect across all mRS levels with the understanding that it may not be constant across all levels. This will be complemented by a graphical assessment of shifts across categories using bar plots as well as binary analyses.

This ordinal analysis will be based on multiple imputations using the same 100 imputed datasets generated for the analysis of the primary outcome. Covariate adjustments are described in Section 4.8.3.3 will also be applied; however, other sensitivity analyses (Section 4.8.3) and subgroup analyses (Section 4.8.4) are not planned.

#### 4.9.2 NIHSS score at 7 days

The NIHSS score at 7 days will be categorized into 7 levels (<5, 5-9, 10-14, 15-19, 20-24, ≥25, and death), and analysed using the same method as for the ordinal analysis of the mRS score described in Section 4.9.1.

The NIHSS score will also be analysed as a continuous variable using a linear regression model similar to the main analysis model (Section 4.8.1), but assuming a normal distribution and an identity link function and without robust/empirical standard errors. The effect of the intervention will be presented as the mean difference and associated 95% CI.

Since we do not expect as much missing data at 7 days as we do at 90 days, the NIHSS analysis will not be based on multiple imputations and will instead use all available data. Covariate adjustments as described in Section 4.8.3.3 will be applied to these analyses of NIHSS; however, other sensitivity analyses (Section 4.8.3) and subgroup analyses (Section 4.8.4) are not planned.

#### 4.9.3 Other binary analyses of mRS at Day 90

Another binary analysis of the mRS at Day 90 will be performed by dichotomizing the mRS as either ‘dependent/dead’ (scores 3-6) or ‘independent’ (scores 0-2) outcomes. This analysis will be conducted using the same approach as the primary analysis described in Section 4.8.1. The effect of the intervention will be presented as the OR of a poor outcome with associated 95% CI. A similar analysis will be performed on mortality alone (mRS of 6) and on dependency alone (mRS 3-5). For dependency alone, the analysis will be restricted to subjects who are alive at Day 90 (mRS 0-5).

These analyses will be based on multiple imputations using the same 100 imputed data sets generated for the analysis of the primary outcome. Covariate adjustments described in Section 4.8.3.3 will also be applied; however, other sensitivity analyses (Section 4.8.3) and subgroup analyses (Section 4.8.4) are not planned.

#### 4.9.4 Investigator-reported sICH according to Heidelberg criteria

The overall proportion of patients who experience an sICH in the intervention and control arms will be reported and compared using logistic regression as per Section 4.8.3.1 but without imputation.

#### 4.9.5 HRQoL

Each of the five EQ-5D dimensions will be described using bar charts. The visual analogous scale ratings will be described using a box plot and summarised as the mean, standard deviation, median and quartiles. The VAS score will be analysed using a linear mixed model using the same approach as for the continuous analysis of NIHSS (see Section 4.9.2). Further analyses of EQ-5D including the heath utility score will be undertaken as part of the economic evaluations, (see Section 4.10).

#### 4.9.6 Time to CT scan

Time from request to CT scan for presumed sICH from last physiological monitoring will be as the mean, standard deviation, median and quartiles, and analysed using a linear mixed model using the same approach as for the continuous analysis of NIHSS (see Section 4.9.2)

#### 4.9.7 Duration of hospitalisation

Duration of hospitalisation will be analysed as time to discharge censored at Day 90 or when the subject was last known to be alive and in hospital, whichever is earlier. It will be summarized using cumulative incidence functions treating mortality as a competing risk. Medians and quartiles of time to discharge will be obtained from the cumulative incidence functions. The effect of the intervention will be estimated as the hazard ratio (HR) (Intervention divided by control) and its 95%CI obtained from a Cox model of the cause-specific hazard which estimates the risk of discharge in subjects who are still alive and have not yet been discharged.[14] Similar to the primary analysis model (Section 4.8.1), the Cox model will include a fixed effect of time as well as a binary variable indicating the group allocation of each patient. To model potential within-cluster correlations, we will use a shared-parameter frailty Cox model with a random site effect.[15]

No imputation, covariate adjustment, subgroup analyses or other sensitivity analysis will be applied to this outcome.

#### 4.9.8 SAEs

SAEs will be summarised as the number of events as well as the number and proportion of patients experiencing at least one event. This will be done overall and by category of event according to Medical Dictionary for Regulatory Activities (MeDRA) system organ classes and preferred terms. The overall proportion of patients with SAEs in the intervention and control arms will be compared using logistic regression as per Section 4.8.3.1 but without imputation. Primary and underlying causes of deaths will be summarised by treatment arm with no formal test.

No imputation, covariate adjustment, subgroup analysis or other sensitivity analysis will be applied to the analysis of SAE. Analyses of SAEs will be run in both the ITT and PP populations (see Section 4.2).

#### 4.9.9 Patient Reported Experience Measurement (PREM) and Sleep-related impairment within 7 days

Reponses to each question will be analysed descriptively using bar charts. For both questionnaires, an overall score will be derived by calculating the average of non-missing items. For the PREM questionnaire, Question 41 (see Figure 2 below) will be excluded from the calculation as it was not always asked. Furthermore, questions 39 and 45 will be set to missing in case the option “Didn’t apply” was selected (Figure 2).

**Figure 2.**
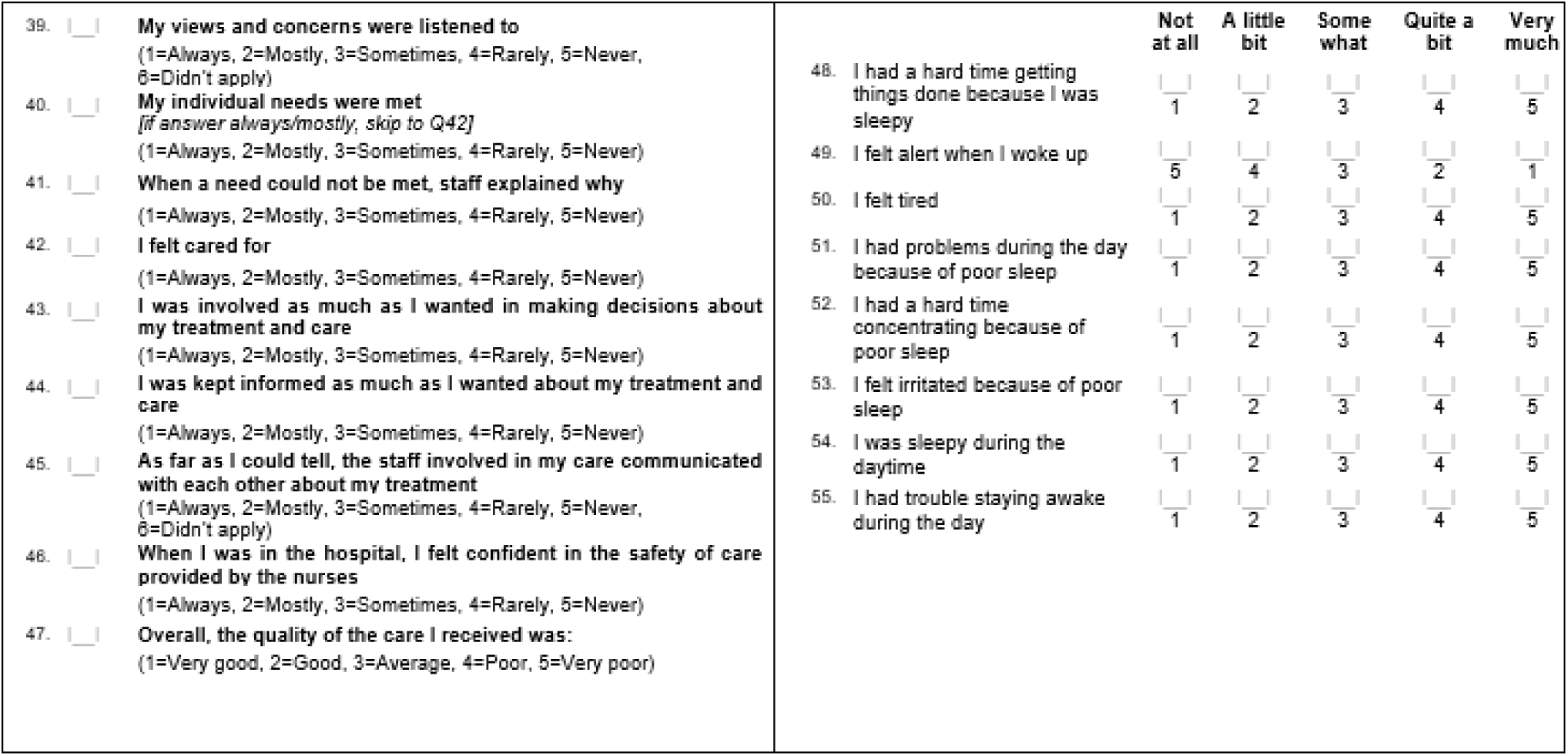
PREM (left) and sleep-related impairment (right) questionnaires

The resulting scores will be summarised using means (SD), and quartiles and analysed using a linear mixed model similar to the one used for the analysis of the NIHSS as a continuous variable (see Section 4.9.2).

### 4.10 Economic and process evaluations

An economic evaluation will be conducted to compare the costs of the standard care and low-intensity monitoring. A cost minimisation analysis will be carried out to determine if the intervention is less expensive (than the standard care) from a healthcare provided perspective

#### 4.10.1 Selection, measurement, and valuation of outcomes

This is a non-inferiority clinical trial where the primary outcome is patient functional recovery defined by an unfavourable outcome (mRS 2-6) at 90 days. With the expectation that the low-intensity monitoring is equally effective, but less resource intensive compared to the standard care, a cost minimisation analysis is carried out with a primary outcome of incremental cost of intervention compared to control group, to determine if the intervention is less expensive (than the standard care).

The costs in this analysis will be estimated from the healthcare provider perspective and reported in 2024 local currency and US dollars. Cost data are collected from the hospital staff survey in the trial or from the existing literature. During patients’ hospitalisation, the cost contributors include personnel, thrombolysis treatment, monitoring consumables, ward, diagnosis, imaging, therapeutics, laboratory analysis and medicine. As the services provided to patients in intervention and control groups only differ in monitoring intensity, the hospitalisation cost only varies in personnel and consumables during the first 24 hours monitoring, and costs for other hospital care components are identical in both groups and thus combined as other cost in the model. Of note, the personnel cost difference during monitoring after thrombolysis treatment is calculated based on its service intensity in the trial. The costs after patients’ discharge consist of medicine, long-term care if required, and re-admission for those having SAE. For the data not available from the trial, costs from existing studies of AIS will be extracted and used in the model.

The economic evaluation will be performed for separate country/region. The cost data will be presented in local currency for each country/region as well as the US dollars in 2024. For data from previous years and from other countries/regions, purchasing power parity will be used to translate the data into the 2024 US dollars and local currencies.

#### 4.10.2 Rationale and description of model

A decision tree will be developed to analyse the costs in intervention and control groups from the perspective of healthcare providers (Figure 3). The model will comprise two arms: the standard care arm and the low-intensity monitoring arm. In each arm, the AIS patients are admitted in the trial hospitals, receive thrombolysis treatment and 24 hours monitoring with different intensities in separate groups. For the patients surviving after 24 hours monitoring, they receive identical hospital care in both groups until discharge. During the 90 day follow up, some of the discharged patients could suffer a SAE.

**Figure 3.**
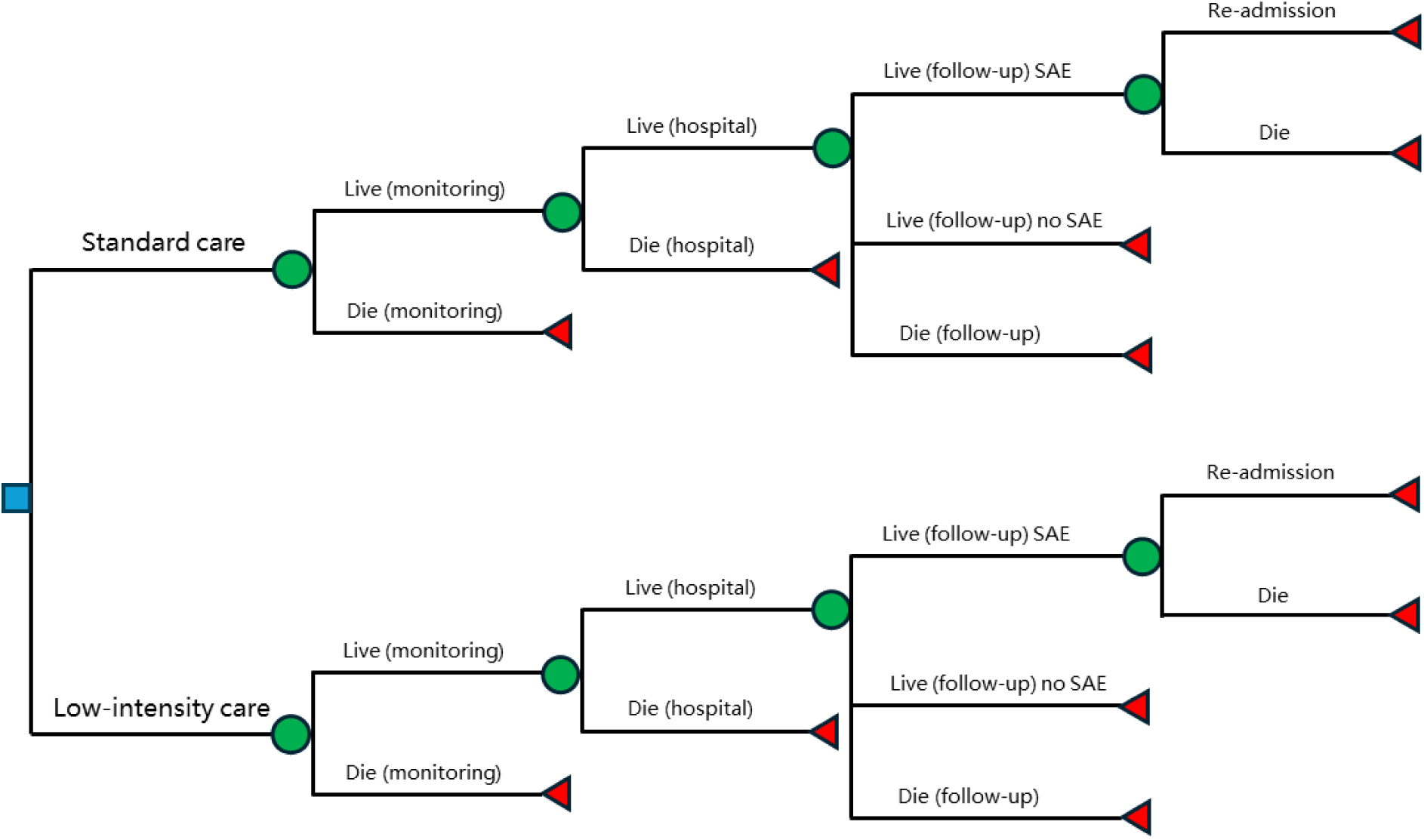
Model structure of cost minimization analysis for OPTIMIST

**Analytics and assumptions:** Patients in the model experience transitions between different states. The transition probabilities will be sourced from the trial and existing studies. Specifically, mortality at 24 hours monitoring, during the admission, and for 90 days of follow-up, and that of SAE occurrence in follow-up period, will be derived from the trial and converted into probabilities as follows:

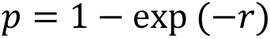

where *p* represents the probabilities and *r* is the rates. Some assumptions will be used in the analysis. First, the characteristics of patients, including their age, sex, and disease severity measured by the baseline NIHSS score, will be assumed to be identical in both groups. Second, due to data unavailability, the other costs in hospital for patients in both groups will be extracted from the data reported from context representative studies and assumed the same in both groups for separate countries/regions. Similarly, the cost for readmission, and probability and cost for long-term care, will also be estimated based on results from country-specific literature. Finally, for the parameters used in the sensitivity analysis without available distribution from literature, a ±50% range will be assumed as the change range.

**Characterising heterogeneity:** First, considering the variance of health system and hospital service in different countries, the economic evaluation will be performed for separate context. Second, subgroup analyses will be conducted for different patient groups regarding their age, sex, and disease severity measured by the baseline NIHSS score.

**Characterising uncertainty:** To assess the influence of model parameter values on the results, sensitivity analyses will be performed. The potential range of the model parameters will be used for the sensitivity analysis. Determination of the change range including the 95% CI of the probabilities and costs from the model, data found from the literature and assumed range of costs.

**Approach to engagement with patients and others affected by the study**: The trial includes an embedded process evaluation which will be separately reported.

**Risks of missing data and potential solutions:**

| Data source | Risks of missing data | Solution |
| --- | --- | --- |
| Data from trial | Cost data missing for a small number of patients in the trial | Data imputation based on patient characteristics (age, sex, disease severity and admitted hospital). |
|  | Cost data missing for all patients from the trial | Replacing the data using the reported costs from published studies conducted in the same patient group. |
| Data from literature | Unavailable country/region representative data for specific trial context | Using data from international studies including the trial country/region or regional studies in the trial country/region. |

## Data Availability

This manuscript is a statistical analysis plan for a trial. The corresponding trial is ongoing and the data is not yet available.

## 6 Proposed outputs

### 6.1 Tables

**Table 1.** Number of subjects enrolled and with primary outcome data per group and per period.

| Group | Randomisation | Period 1 | Period 2 | Period 3 | Total |
| --- | --- | --- | --- | --- | --- |
| **1 (xx sites)** | xxx / xxx (xx%) | xxx / xxx (xx%) | xxx / xxx (xx%) | xxx / xxx (xx%) | xxx / xxx (xx%) |
| **2 (xx sites)** | xxx / xxx (xx%) | xxx / xxx (xx%) | xxx / xxx (xx%) | xxx / xxx (xx%) | xxx / xxx (xx%) |
| **3 (xx sites)** | xxx / xxx (xx%) | xxx / xxx (xx%) | xxx / xxx (xx%) | xxx / xxx (xx%) | xxx / xxx (xx%) |
| **Total (xxx sites)** | xxx / xxx (xx%) | xxx / xxx (xx%) | xxx / xxx (xx%) | xxx / xxx (xx%) | xxx / xxx (xx%) |

Notes: the denominator represents the number of subjects enrolled during the period. The numerator represents the number of subjects with a primary endpoint measure (mRS at Day 90). Cells shaded in grey represent usual care periods. Cells shaded in orange represent care bundle periods.

**Table 2.** Baseline characteristics.

|  | Low intensity<br>(N = ) | Standard care<br>(N = ) | Total<br>(N = ) |
| --- | --- | --- | --- |
| **Age (years)** | xxx | xxx | xxx |
| Mean (SD) | xxx.x (xxx.x) | xxx.x (xxx.x) | xxx.x (xxx.x) |
| Median (Q1; Q3) | xxx (xxx; xxx) | xxx (xxx; xxx) | xxx (xxx; xxx) |
| min max | xxx to xxx | xxx to xxx | xxx to xxx |
| **Sex** | xxx | xxx | xxx |
| Male | xxx xx.x% | xxx xx.x% | xxx xx.x% |
| Female | xxx xx.x% | xxx xx.x% | xxx xx.x% |
| **Ethnicity** | xxx | xxx | xxx |
| Black (include African American; of African origins) | xxx xx.x% | xxx xx.x% | xxx xx.x% |
| Asian (include Far East, South East Asian and Indian subcontinent) | xxx xx.x% | xxx xx.x% | xxx xx.x% |
| Hispanic (include Latin America, Spanish) | xxx xx.x% | xxx xx.x% | xxx xx.x% |
| White/Caucasian (include European, Middle East, North Africa) | xxx xx.x% | xxx xx.x% | xxx xx.x% |
| Indigenous (include Aboriginal, Pacific Islander, Native Hawaiian) | xxx xx.x% | xxx xx.x% | xxx xx.x% |
| Uncertain | xxx xx.x% | xxx xx.x% | xxx xx.x% |
| Other | xxx xx.x% | xxx xx.x% | xxx xx.x% |
| **Current smoker** | xxx xx.x% | xxx xx.x% | xxx xx.x% |
| **SBP (mmHg)** | xxx | xxx | xxx |
| Mean (SD) | xxx.x (xxx.x) | xxx.x (xxx.x) | xxx.x (xxx.x) |
| Median (Q1; Q3) | xxx (xxx; xxx) | xxx (xxx; xxx) | xxx (xxx; xxx) |
| min max | xxx to xxx | xxx to xxx | xxx to xxx |
| **DBP (mmHg)** | xxx | xxx | xxx |
| Mean (SD) | xxx.x (xxx.x) | xxx.x (xxx.x) | xxx.x (xxx.x) |
| Median (Q1; Q3) | xxx (xxx; xxx) | xxx (xxx; xxx) | xxx (xxx; xxx) |
| min max | xxx to xxx | xxx to xxx | xxx to xxx |
| **Heart Rate (beats/min)** | xxx | xxx | xxx |
| Mean (SD) | xxx.x (xxx.x) | xxx.x (xxx.x) | xxx.x (xxx.x) |
| Median (Q1; Q3) | xxx (xxx; xxx) | xxx (xxx; xxx) | xxx (xxx; xxx) |
| min max | xxx to xxx | xxx to xxx | xxx to xxx |
| **GCS score** | xxx | xxx | xxx |
| Mean (SD) | xxx.x (xxx.x) | xxx.x (xxx.x) | xxx.x (xxx.x) |
| Median (Q1; Q3) | xxx (xxx; xxx) | xxx (xxx; xxx) | xxx (xxx; xxx) |
| min max | xxx to xxx | xxx to xxx | xxx to xxx |
| **NIHSS score on arrival** | xxx | xxx | xxx |
| Mean (SD) | xxx.x (xxx.x) | xxx.x (xxx.x) | xxx.x (xxx.x) |
| Median (Q1; Q3) | xxx (xxx; xxx) | xxx (xxx; xxx) | xxx (xxx; xxx) |
| min max | xxx to xxx | xxx to xxx | xxx to xxx |
| **NIHSS score after thrombolysis** | xxx | xxx | xxx |
| Mean (SD) | xxx.x (xxx.x) | xxx.x (xxx.x) | xxx.x (xxx.x) |
| Median (Q1; Q3) | xxx (xxx; xxx) | xxx (xxx; xxx) | xxx (xxx; xxx) |
| min max | xxx to xxx | xxx to xxx | xxx to xxx |
| **Undertaking endovascular treatment** |  |  |  |
| No | xxx xx.x% | xxx xx.x% | xxx xx.x% |
| Yes | xxx xx.x% | xxx xx.x% | xxx xx.x% |
| **Modified Treatment in Cerebral Ischaemia score (mTICI)** |  |  |  |
| Grade 0 - no perfusion | xxx.x (xxx.x) | xxx.x (xxx.x) | xxx.x (xxx.x) |
| Grade 2b - partial perfusion 50-99% | xxx.x (xxx.x) | xxx.x (xxx.x) | xxx.x (xxx.x) |
| Grade 3 - complete perfusion | xxx.x (xxx.x) | xxx.x (xxx.x) | xxx.x (xxx.x) |
| Uncertain | xxx.x (xxx.x) | xxx.x (xxx.x) | xxx.x (xxx.x) |
| Not applicable - did not have endovascular treatment | xxx.x (xxx.x) | xxx.x (xxx.x) | xxx.x (xxx.x) |
| **Neurological status** |  |  |  |
| Stable | xxx.x (xxx.x) | xxx.x (xxx.x) | xxx.x (xxx.x) |
| Improving | xxx.x (xxx.x) | xxx.x (xxx.x) | xxx.x (xxx.x) |
| Worse but not requiring critical care | xxx.x (xxx.x) | xxx.x (xxx.x) | xxx.x (xxx.x) |
| Worse and requiring critical care | xxx.x (xxx.x) | xxx.x (xxx.x) | xxx.x (xxx.x) |
| **Pre-morbid function on the modified Rankin scale** | xxx | xxx | xxx |
| 0 - no symptoms | xxx xx.x% | xxx xx.x% | xxx xx.x% |
| 1 - no significant disability (with symptoms) | xxx xx.x% | xxx xx.x% | xxx xx.x% |
| 2 - slight disability (but independent in daily activities) | xxx xx.x% | xxx xx.x% | xxx xx.x% |
| 3 - moderate disability (requiring some help from another person for daily activities) | xxx xx.x% | xxx xx.x% | xxx xx.x% |
| 4 - moderate severe disability (requiring regular help from another person) | xxx xx.x% | xxx xx.x% | xxx xx.x% |
| 5 - severe disability (bed bound, totally dependent) | xxx xx.x% | xxx xx.x% | xxx xx.x% |

**Table 3.** Medical history.

| Variable | Low intensity<br>(N = ) | Standard care<br>(N = ) | Total<br>(N = ) |
| --- | --- | --- | --- |
| Previous ischaemic stroke | xxx/xxx xx.x% | xxx/xxx xx.x% | xxx/xxx xx.x% |
| Previous intracerebral haemorrhage | xxx/xxx xx.x% | xxx/xxx xx.x% | xxx/xxx xx.x% |
| Coronary heart disease | xxx/xxx xx.x% | xxx/xxx xx.x% | xxx/xxx xx.x% |
| Documented atrial fibrillation | xxx/xxx xx.x% | xxx/xxx xx.x% | xxx/xxx xx.x% |
| Other heart disease | xxx/xxx xx.x% | xxx/xxx xx.x% | xxx/xxx xx.x% |
| Hypertension | xxx/xxx xx.x% | xxx/xxx xx.x% | xxx/xxx xx.x% |
| Diabetes | xxx/xxx xx.x% | xxx/xxx xx.x% | xxx/xxx xx.x% |
| Hypercholesterolaemia | xxx/xxx xx.x% | xxx/xxx xx.x% | xxx/xxx xx.x% |

**Table 4.** Medications at time of admission.

| Variable | Low intensity<br>(N = ) | Standard care<br>(N = ) | Total<br>(N = ) |
| --- | --- | --- | --- |
| Antihypertensive agent(s) | xxx/xxx xx.x% | xxx/xxx xx.x% | xxx/xxx xx.x% |
| Anticoagulant | xxx/xxx xx.x% | xxx/xxx xx.x% | xxx/xxx xx.x% |
| Anti-platelet agent | xxx/xxx xx.x% | xxx/xxx xx.x% | xxx/xxx xx.x% |
| Glucose lowering treatment | xxx/xxx xx.x% | xxx/xxx xx.x% | xxx/xxx xx.x% |
| Lipid lowering agent(s) | xxx/xxx xx.x% | xxx/xxx xx.x% | xxx/xxx xx.x% |

**Table 5.** Vital signs recorded at Day 1 (first 24 hours)

| Variable | Low intensity<br>(N = ) | Standard care<br>(N = ) | Total<br>(N = ) |
| --- | --- | --- | --- |
| **SBP (mmHg)** |  |  |  |
| Mean (SD) | xxx.x (xxx.x) | xxx.x (xxx.x) | xxx.x (xxx.x) |
| Median (Q1; Q3) | xxx (xxx; xxx) | xxx (xxx; xxx) | xxx (xxx; xxx) |
| min max | xxx to xxx | xxx to xxx | xxx to xxx |
| **DBP (mmHg)** |  |  |  |
| Mean (SD) | xxx.x (xxx.x) | xxx.x (xxx.x) | xxx.x (xxx.x) |
| Median (Q1; Q3) | xxx (xxx; xxx) | xxx (xxx; xxx) | xxx (xxx; xxx) |
| min max | xxx to xxx | xxx to xxx | xxx to xxx |
| **Heart rate (bpm)** | xxx | xxx | xxx |
| Mean (SD) | xxx.x (xxx.x) | xxx.x (xxx.x) | xxx.x (xxx.x) |
| Median (Q1; Q3) | xxx (xxx; xxx) | xxx (xxx; xxx) | xxx (xxx; xxx) |
| min max | xxx to xxx | xxx to xxx | xxx to xxx |

**Table 6.** Management and treatments during the first 24 hours.

| Variable | Low intensity<br>(N = ) | Standard care<br>(N = ) | Total<br>(N = ) |
| --- | --- | --- | --- |
| **Allocated post-thrombolysis monitoring protocol followed** | xxx/xxx xx.x% | xxx/xxx xx.x% | xxx/xxx xx.x% |
| **Duration of monitoring (hours)** | xxx | xxx | xxx |
| Mean (SD) | xxx.x (xxx.x) | xxx.x (xxx.x) | xxx.x (xxx.x) |
| Median (Q1; Q3) | xxx (xxx; xxx) | xxx (xxx; xxx) | xxx (xxx; xxx) |
| min max | xxx to xxx | xxx to xxx | xxx to xxx |
| **Neurological assessments (GCS and/or NIHSS) taken during monitoring care** | xxx | xxx | xxx |
| Mean (SD) | xxx.x (xxx.x) | xxx.x (xxx.x) | xxx.x (xxx.x) |
| Median (Q1; Q3) | xxx (xxx; xxx) | xxx (xxx; xxx) | xxx (xxx; xxx) |
| min max | xxx to xxx | xxx to xxx | xxx to xxx |
| **Vital signs (HR and BP) measurements taken during monitoring care** |  |  |  |
| Mean (SD) | xxx.x (xxx.x) | xxx.x (xxx.x) | xxx.x (xxx.x) |
| Median (Q1; Q3) | xxx (xxx; xxx) | xxx (xxx; xxx) | xxx (xxx; xxx) |
| min max | xxx to xxx | xxx to xxx | xxx to xxx |
| **Intravenous thrombolytic agent used** |  |  |  |
| Alteplase | xxx/xxx xx.x% | xxx/xxx xx.x% | xxx/xxx xx.x% |
| Tenecteplase | xxx/xxx xx.x% | xxx/xxx xx.x% | xxx/xxx xx.x% |
| Other | xxx/xxx xx.x% | xxx/xxx xx.x% | xxx/xxx xx.x% |
| **Alteplase dose (mg)** | xxx | xxx | xxx |
| Mean (SD) | xxx.x (xxx.x) | xxx.x (xxx.x) | xxx.x (xxx.x) |
| Median (Q1; Q3) | xxx (xxx; xxx) | xxx (xxx; xxx) | xxx (xxx; xxx) |
| min max | xxx to xxx | xxx to xxx | xxx to xxx |
| Mean (SD) | xxx.x (xxx.x) | xxx.x (xxx.x) | xxx.x (xxx.x) |
| **Bolus dose (mg)** | xxx | xxx | xxx |
| Mean (SD) | xxx.x (xxx.x) | xxx.x (xxx.x) | xxx.x (xxx.x) |
| Median (Q1; Q3) | xxx (xxx; xxx) | xxx (xxx; xxx) | xxx (xxx; xxx) |
| min max | xxx to xxx | xxx to xxx | xxx to xxx |
| Mean (SD) | xxx.x (xxx.x) | xxx.x (xxx.x) | xxx.x (xxx.x) |
| **Location of immediate post-thrombolysis care** |  |  |  |
| Acute Stroke Unit | xxx/xxx xx.x% | xxx/xxx xx.x% | xxx/xxx xx.x% |
| Intensive Care Unit | xxx/xxx xx.x% | xxx/xxx xx.x% | xxx/xxx xx.x% |
| Intermediate Care Unit (IMC) / Step Down Unit or<br>High Dependency Care Unit | xxx/xxx xx.x% | xxx/xxx xx.x% | xxx/xxx xx.x% |
| Other | xxx/xxx xx.x% | xxx/xxx xx.x% | xxx/xxx xx.x% |
| **Neurologically stable or improved** |  |  |  |
| No | xxx/xxx xx.x% | xxx/xxx xx.x% | xxx/xxx xx.x% |
| Yes | xxx/xxx xx.x% | xxx/xxx xx.x% | xxx/xxx xx.x% |
| **Change to monitored care in first 24 hours** |  |  |  |
| Yes | xxx/xxx xx.x% | xxx/xxx xx.x% | xxx/xxx xx.x% |
| No | xxx/xxx xx.x% | xxx/xxx xx.x% | xxx/xxx xx.x% |
| **Reason for interruption of the monitoring care** |  |  |  |
| Medical event | xxx/xxx xx.x% | xxx/xxx xx.x% | xxx/xxx xx.x% |
| Nursing preference | xxx/xxx xx.x% | xxx/xxx xx.x% | xxx/xxx xx.x% |
| Neurological deterioration | xxx/xxx xx.x% | xxx/xxx xx.x% | xxx/xxx xx.x% |
| Other | xxx/xxx xx.x% | xxx/xxx xx.x% | xxx/xxx xx.x% |

**Table 7.** Diagnosis at Day 7 and management/treatments received during hospitalisation.

| Variable | Low intensity<br>(N = ) | Standard care<br>(N = ) | Total<br>(N = ) |
| --- | --- | --- | --- |
| **Patient status** |  |  |  |
| Still in Hospital | xxx/xxx xx.x% | xxx/xxx xx.x% | xxx/xxx xx.x% |
| Discharge or transfer | xxx/xxx xx.x% | xxx/xxx xx.x% | xxx/xxx xx.x% |
| Died | xxx/xxx xx.x% | xxx/xxx xx.x% | xxx/xxx xx.x% |
| **Final diagnosis was an acute ischaemic stroke** |  |  |  |
| No | xxx/xxx xx.x% | xxx/xxx xx.x% | xxx/xxx xx.x% |
| Yes | xxx/xxx xx.x% | xxx/xxx xx.x% | xxx/xxx xx.x% |
| **Non-stroke diagnosis** |  |  |  |
| Migraine | xxx/xxx xx.x% | xxx/xxx xx.x% | xxx/xxx xx.x% |
| Seizure | xxx/xxx xx.x% | xxx/xxx xx.x% | xxx/xxx xx.x% |
| Functional weakness | xxx/xxx xx.x% | xxx/xxx xx.x% | xxx/xxx xx.x% |
| Syncope | xxx/xxx xx.x% | xxx/xxx xx.x% | xxx/xxx xx.x% |
| Other | xxx/xxx xx.x% | xxx/xxx xx.x% | xxx/xxx xx.x% |
| **Final stroke diagnosis** |  |  |  |
| Large vessel atheroma (>50% stenosis) in extra- or intra-cranial vessel | xxx/xxx xx.x% | xxx/xxx xx.x% | xxx/xxx xx.x% |
| Small vessel or perforating vessel lacunar disease | xxx/xxx xx.x% | xxx/xxx xx.x% | xxx/xxx xx.x% |
| Cardio-emboli | xxx/xxx xx.x% | xxx/xxx xx.x% | xxx/xxx xx.x% |
| Dissection | xxx/xxx xx.x% | xxx/xxx xx.x% | xxx/xxx xx.x% |
| Uncertain aetiology | xxx/xxx xx.x% | xxx/xxx xx.x% | xxx/xxx xx.x% |
| Other definite pathological mechanism | xxx/xxx xx.x% | xxx/xxx xx.x% | xxx/xxx xx.x% |
| **Oxfordshire Community Stroke Project (OCSP) classification** |  |  |  |
| Total Anterior Circulation Infarct (TACI) | xxx/xxx xx.x% | xxx/xxx xx.x% | xxx/xxx xx.x% |
| Partial Anterior Circulation Infarct (PACI) | xxx/xxx xx.x% | xxx/xxx xx.x% | xxx/xxx xx.x% |
| Lacunar Infarct (LACI) | xxx/xxx xx.x% | xxx/xxx xx.x% | xxx/xxx xx.x% |
| Posterior Circulation infarct (POCI) | xxx/xxx xx.x% | xxx/xxx xx.x% | xxx/xxx xx.x% |
|  | xxx/xxx xx.x% | xxx/xxx xx.x% | xxx/xxx xx.x% |
| **Treatment during hospitalization** | xxx/xxx xx.x% | xxx/xxx xx.x% | xxx/xxx xx.x% |
| Intubation and ventilation | xxx/xxx xx.x% | xxx/xxx xx.x% | xxx/xxx xx.x% |
| Tracheostomy | xxx/xxx xx.x% | xxx/xxx xx.x% | xxx/xxx xx.x% |
| Nasogastric feeding given | xxx/xxx xx.x% | xxx/xxx xx.x% | xxx/xxx xx.x% |
| Gastrostomy tube inserted | xxx/xxx xx.x% | xxx/xxx xx.x% | xxx/xxx xx.x% |
| Patient mobilised by physiotherapist/occupational therapist | xxx/xxx xx.x% | xxx/xxx xx.x% | xxx/xxx xx.x% |
| Compression cuffs used/mechanical (TED) | xxx/xxx xx.x% | xxx/xxx xx.x% | xxx/xxx xx.x% |
| Subcutaneous heparin or heparinoids administered | xxx/xxx xx.x% | xxx/xxx xx.x% | xxx/xxx xx.x% |
| Glycaemic control given | xxx/xxx xx.x% | xxx/xxx xx.x% | xxx/xxx xx.x% |
| Intravenous heparin administered | xxx/xxx xx.x% | xxx/xxx xx.x% | xxx/xxx xx.x% |
| Aspirin/other anti-platelet agent administered | xxx/xxx xx.x% | xxx/xxx xx.x% | xxx/xxx xx.x% |
| Intravenous steroids administered | xxx/xxx xx.x% | xxx/xxx xx.x% | xxx/xxx xx.x% |
| Hemicraniectomy or suboccipital craniectomy performed | xxx/xxx xx.x% | xxx/xxx xx.x% | xxx/xxx xx.x% |
| Acute Stroke Unit admission | xxx/xxx xx.x% | xxx/xxx xx.x% | xxx/xxx xx.x% |
| Intensive Care Unit admission | xxx/xxx xx.x% | xxx/xxx xx.x% | xxx/xxx xx.x% |
| IMC/stepdown admission | xxx/xxx xx.x% | xxx/xxx xx.x% | xxx/xxx xx.x% |
| Requirement for any form of renal dialysis | xxx/xxx xx.x% | xxx/xxx xx.x% | xxx/xxx xx.x% |
| Clinical decision made to withdraw 'active' care for the patient | xxx/xxx xx.x% | xxx/xxx xx.x% | xxx/xxx xx.x% |
| Undertaken formal rehabilitation assessment | xxx/xxx xx.x% | xxx/xxx xx.x% | xxx/xxx xx.x% |

**Table 8.** Hospital readmission and health service use.

| Variable | Low intensity<br>(N = ) | Standard care<br>(N = ) | Total<br>(N = ) |
| --- | --- | --- | --- |
| **Hospital re-admission** | xxx | xxx | xxx |
| Mean (SD) | xxx.x (xxx.x) | xxx.x (xxx.x) | xxx.x (xxx.x) |
| Median (Q1; Q3) | xxx (xxx; xxx) | xxx (xxx; xxx) | xxx (xxx; xxx) |
| min max | xxx to xxx | xxx to xxx | xxx to xxx |
| **Doctors or health professional visits** | xxx | xxx | xxx |
| Mean (SD) | xxx.x (xxx.x) | xxx.x (xxx.x) | xxx.x (xxx.x) |
| Median (Q1; Q3) | xxx (xxx; xxx) | xxx (xxx; xxx) | xxx (xxx; xxx) |
| min max | xxx to xxx | xxx to xxx | xxx to xxx |

**Table 9.** Clinical outcomes - Descriptive analysis.

| Variable | Low intensity<br>(N = ) | Standard care<br>(N = ) | Total<br>(N = ) |
| --- | --- | --- | --- |
| **NIHSS** | xxx | xxx | xxx |
| **Day 1** |  |  |  |
| Mean (SD) | xxx.x (xxx.x) | xxx.x (xxx.x) | xxx.x (xxx.x) |
| Median (Q1; Q3) | xxx (xxx; xxx) | xxx (xxx; xxx) | xxx (xxx; xxx) |
| min max | xxx to xxx | xxx to xxx | xxx to xxx |
| **Day 7** |  |  |  |
| Mean (SD) | xxx.x (xxx.x) | xxx.x (xxx.x) | xxx.x (xxx.x) |
| Median (Q1; Q3) | xxx (xxx; xxx) | xxx (xxx; xxx) | xxx (xxx; xxx) |
| min max | xxx to xxx | xxx to xxx | xxx to xxx |
| **Categorical scores on the NIHSS or death at 7 days** |  |  |  |
| Scores 0–4 | xxx/xxx xx.x% | xxx/xxx xx.x% | xxx/xxx xx.x% |
| Scores 5–9 | xxx/xxx xx.x% | xxx/xxx xx.x% | xxx/xxx xx.x% |
| Scores 10–14 | xxx/xxx xx.x% | xxx/xxx xx.x% | xxx/xxx xx.x% |
| Scores 15–19 | xxx/xxx xx.x% | xxx/xxx xx.x% | xxx/xxx xx.x% |
| Scores 20–24 | xxx/xxx xx.x% | xxx/xxx xx.x% | xxx/xxx xx.x% |
| Scores ≥25 | xxx/xxx xx.x% | xxx/xxx xx.x% | xxx/xxx xx.x% |
| Death | xxx/xxx xx.x% | xxx/xxx xx.x% | xxx/xxx xx.x% |
| **mRS score** |  |  |  |
| **Day 7** |  |  |  |
| 0 - no symptoms | xxx/xxx xx.x% | xxx/xxx xx.x% | xxx/xxx xx.x% |
| 1 - no significant disability | xxx/xxx xx.x% | xxx/xxx xx.x% | xxx/xxx xx.x% |
| 2 - slight disability | xxx/xxx xx.x% | xxx/xxx xx.x% | xxx/xxx xx.x% |
| 3 - moderate disability | xxx/xxx xx.x% | xxx/xxx xx.x% | xxx/xxx xx.x% |
| 4 - moderate severe disability | xxx/xxx xx.x% | xxx/xxx xx.x% | xxx/xxx xx.x% |
| 5 - severe disability | xxx/xxx xx.x% | xxx/xxx xx.x% | xxx/xxx xx.x% |
| 6 - Death | xxx/xxx xx.x% | xxx/xxx xx.x% | xxx/xxx xx.x% |
| **Day 90** |  |  |  |
| 0 - no symptoms | xxx/xxx xx.x% | xxx/xxx xx.x% | xxx/xxx xx.x% |
| 1 - no significant disability | xxx/xxx xx.x% | xxx/xxx xx.x% | xxx/xxx xx.x% |
| 2 - slight disability | xxx/xxx xx.x% | xxx/xxx xx.x% | xxx/xxx xx.x% |
| 3 - moderate disability | xxx/xxx xx.x% | xxx/xxx xx.x% | xxx/xxx xx.x% |
| 4 - moderate severe disability | xxx/xxx xx.x% | xxx/xxx xx.x% | xxx/xxx xx.x% |
| 5 - severe disability | xxx/xxx xx.x% | xxx/xxx xx.x% | xxx/xxx xx.x% |
| 6 - Death | xxx/xxx xx.x% | xxx/xxx xx.x% | xxx/xxx xx.x% |
| **Bad outcome (mRS 2-6 vs 0-1)** | xxx/xxx xx.x% | xxx/xxx xx.x% | xxx/xxx xx.x% |
| **Death or major disability (mRS 3-6 vs 0-2)** | xxx/xxx xx.x% | xxx/xxx xx.x% | xxx/xxx xx.x% |
| **All-cause mortality (mRS 6 vs 0-5)** | xxx/xxx xx.x% | xxx/xxx xx.x% | xxx/xxx xx.x% |
| **Major disability (mRS 3-5 vs 0-2)** | xxx/xxx xx.x% | xxx/xxx xx.x% | xxx/xxx xx.x% |
| **Investigator-reported sICH according to Heidelberg criteria** | xxx/xxx xx.x% | xxx/xxx xx.x% | xxx/xxx xx.x% |
| **Time to CT scan** |  |  |  |
| Mean (SD) | xxx.x (xxx.x) | xxx.x (xxx.x) | xxx.x (xxx.x) |
| Median (Q1; Q3) | xxx (xxx; xxx) | xxx (xxx; xxx) | xxx (xxx; xxx) |
| min max | xxx to xxx | xxx to xxx | xxx to xxx |
| **EQ-5D VAS score at 90 days** |  |  |  |
| Mean (SD) | xxx.x (xxx.x) | xxx.x (xxx.x) | xxx.x (xxx.x) |
| Median (Q1; Q3) | xxx (xxx; xxx) | xxx (xxx; xxx) | xxx (xxx; xxx) |
| min max | xxx to xxx | xxx to xxx | xxx to xxx |
| **PREM at 7 days** |  |  |  |
| Mean (SD) | xxx.x (xxx.x) | xxx.x (xxx.x) | xxx.x (xxx.x) |
| Median (Q1; Q3) | xxx (xxx; xxx) | xxx (xxx; xxx) | xxx (xxx; xxx) |
| min max | xxx to xxx | xxx to xxx | xxx to xxx |
| **Sleep-related impairment at 7 days** |  |  |  |
| Mean (SD) | xxx.x (xxx.x) | xxx.x (xxx.x) | xxx.x (xxx.x) |
| Median (Q1; Q3) | xxx (xxx; xxx) | xxx (xxx; xxx) | xxx (xxx; xxx) |
| min max | xxx to xxx | xxx to xxx | xxx to xxx |

**Table 10.** Clinical outcomes – main model results.

| Outcome [nature] | Unadjusted model |  |  | Adjusted model |  |  |
| --- | --- | --- | --- | --- | --- | --- |
|  | N | Estimate (95% CI) | P | N | Estimate (95% CI) | P |
| **Bad outcome (mRS 2-6 vs 0-1) at Day 90 [binary]** | xxxx | xx.xx (xx.xx to xx.xx) | **0.xxx** | xxxx | xx.xx (xx.xx to xx.xx) | **0.xxx** |
| Shift analysis of mRS at Day 90 [ordinal] | xxxx | xx.xx (xx.xx to xx.xx) |  | xxxx | xx.xx (xx.xx to xx.xx) |  |
| NIHSS score at 7 days [ordinal] | xxxx | xx.xx (xx.xx to xx.xx) |  | xxxx | xx.xx (xx.xx to xx.xx) |  |
| NIHSS score at 7 days [continuous] | xxxx | xx.xx (xx.xx to xx.xx) |  | xxxx | xx.xx (xx.xx to xx.xx) |  |
| Death or dependency (mRS 3-6 vs 0-2) at Day 90 [binary] | xxxx | xx.xx (xx.xx to xx.xx) |  | xxxx | xx.xx (xx.xx to xx.xx) |  |
| Death (mRS 6 vs 0-5) at Day 90 [binary] | xxxx | xx.xx (xx.xx to xx.xx) |  | xxxx | xx.xx (xx.xx to xx.xx) |  |
| Dependency (mRS 2-5 vs 0-2) at Day 90 [binary] | xxxx | xx.xx (xx.xx to xx.xx) |  | xxxx | xx.xx (xx.xx to xx.xx) |  |
| Investigator-reported sICH [binary] | xxxx | xx.xx (xx.xx to xx.xx) |  | xxxx | xx.xx (xx.xx to xx.xx) |  |
| Time to CT scan [continuous] | xxxx | xx.xx (xx.xx to xx.xx) |  | xxxx | xx.xx (xx.xx to xx.xx) |  |
| Time to hospital discharge [survival] | xxxx | xx.xx (xx.xx to xx.xx) |  | xxxx | xx.xx (xx.xx to xx.xx) |  |
| EQ-5D VAS score at 90 days [continuous] | xxxx | xx.xx (xx.xx to xx.xx) |  | xxxx | xx.xx (xx.xx to xx.xx) |  |
| PREM at 7 days [continuous] | xxxx | xx.xx (xx.xx to xx.xx) |  | xxxx | xx.xx (xx.xx to xx.xx) |  |
| Sleep-related impairment at 7 days [continuous] | xxxx | xx.xx (xx.xx to xx.xx) |  | xxxx | xx.xx (xx.xx to xx.xx) |  |

- 1) For ordinal, continuous and binary outcomes, the unadjusted model consists in a generalised linear mixed model with a random effect for cluster (hospital site), a fixed effect indicating the group assignment of each cluster at each step and a fixed categorical effect of time. Appropriate distributions and link functions are used depending on the nature of the outcome. For survival outcomes, we use a Cox model with a random cluster effect. - 2) Adjusted models include the following baseline covariates: Country, Baseline mRS, Age, Sex, Baseline NIHSS score - 3) All models are adjusted for time using calendar time in 6-month intervals - 4) All analyses of mRS presented in this table are based on data imputed using multiple imputations under a MAR assumption. Analyses of other outcomes are based on all data available (no imputation) - 5) For the primary outcome, the p-value corresponds to a test of non-inferiority to reject a RR of 1.15. For secondary outcomes, we focus on point and interval estimation rather than significance testing given that we do not expect true differences between the two arms - 6) For dependency alone, the analysis will be restricted to subjects who are alive at Day 90 (mRS 0-5)

**Table 11.** Sensitivity analyses of the primary outcome.

| Model / Modelling of time / Missing data handling | Unadjusted model <sup>1</sup> |  |  | Adjusted model <sup>2</sup> |  |  |
| --- | --- | --- | --- | --- | --- | --- |
|  | N | OR/MD/HR (95% CI) | P | N | OR/MD/HR (95% CI) | P |
| **Log-binomial regression** |  |  |  |  |  |  |
| Calendar time (6 months) with common time trend to all clusters <sup>3</sup> |  |  |  |  |  |  |
| *Multiple imputations (MAR) <sup>5</sup>* | xxxx | xx.xx (xx.xx to xx.xx) | 0.xxx | xxxx | xx.xx (xx.xx to xx.xx) | 0.xxx |
| *Using available data* | xxxx | xx.xx (xx.xx to xx.xx) | 0.xxx | xxxx | xx.xx (xx.xx to xx.xx) | 0.xxx |
| *Multiple imputations (MNAR) <sup>7</sup>* | xxxx | xx.xx (xx.xx to xx.xx) | 0.xxx | xxxx | xx.xx (xx.xx to xx.xx) | 0.xxx |
| Calendar time (6 months) allowing time trends to vary by cluster (MAR) <sup>4</sup> | xxxx | xx.xx (xx.xx to xx.xx) | 0.xxx | xxxx | xx.xx (xx.xx to xx.xx) | 0.xxx |
| Restricted Cubic splines with calendar time in 1-month intervals (MAR) | xxxx | xx.xx (xx.xx to xx.xx) | 0.xxx | xxxx | xx.xx (xx.xx to xx.xx) | 0.xxx |
| Time as stepped-wedge period (MAR) | xxxx | xx.xx (xx.xx to xx.xx) | 0.xxx | xxxx | xx.xx (xx.xx to xx.xx) | 0.xxx |
| **Logistic regression** |  |  |  |  |  |  |
| Calendar time (6 months) with common time trend to all clusters (MAR) <sup>3</sup> | xxxx | xx.xx (xx.xx to xx.xx) | n/a | xxxx | xx.xx (xx.xx to xx.xx) | n/a |

Note: this table will be repeated in the per-protocol set (see Section 4.2 for definitions)

**Table 12.** SAE.

| Event | Intervention<br>(N = ) | Usual Care<br>(N = ) | P-value |
| --- | --- | --- | --- |
| **Any SAE** | **nEVT nPT(xx.x%)** | **nEVT nPT(xx.x%)** | **0.xx** |
| Resulted in death | nEVT nPT(xx.x%) | nEVT nPT(xx.x%) |  |
| Life threatening | nEVT nPT(xx.x%) | nEVT nPT(xx.x%) |  |
| Requires prolonged hospitalisation | nEVT nPT(xx.x%) | nEVT nPT(xx.x%) |  |
| Results in persistent or severe disability/incapacity | nEVT nPT(xx.x%) | nEVT nPT(xx.x%) |  |
| Results in congenital anomaly/birth defect | nEVT nPT(xx.x%) | nEVT nPT(xx.x%) |  |
| Medically significant | nEVT nPT(xx.x%) | nEVT nPT(xx.x%) |  |
| **Days from enrolment to SAE onset** | xxx | xxx |  |
| Mean (SD) | xxx.x (xxx.x) | xxx.x (xxx.x) |  |
| Median (Q1; Q3) | xxx (xxx; xxx) | xxx (xxx; xxx) |  |
| min max | xxx to xxx | xxx to xxx |  |
| **Investigator-reported symptomatic ICH** |  |  |  |
| Associated with major neurological deterioration | nEVT nPT(xx.x%) | nEVT nPT(xx.x%) |  |
| Associated with minor neurological deterioration | nEVT nPT(xx.x%) | nEVT nPT(xx.x%) |  |
| **Events by system/disease** |  |  |  |
| Cardiac | nEVT nPT(xx.x%) | nEVT nPT(xx.x%) |  |
| Nervous system | nEVT nPT(xx.x%) | nEVT nPT(xx.x%) |  |
| Etc... | nEVT nPT(xx.x%) | nEVT nPT(xx.x%) |  |

*Programming note:*
*AEs will be summarised as the number (nPT) and proportion of patients experiencing at least one event. In addition, the total number of events (nEVT) will be reported.*
*AEs will be manually allocated to broad system/disease categories*
Note: this table will be repeated in the per-protocol set (see Section 3.2 for definitions)

**Table 13.** Causes of death.

| Cause | Intervention<br>(N = ) | Usual Care<br>(N = ) |
| --- | --- | --- |
| **Proximate cause of death** | N=XXX | N=XXX |
| Most common cause #1 | nnn xx% | nnn xx% |
| Most common cause #2 | nnn xx% | nnn xx% |
| Etc. |  |  |
| Most common cause #10 | nnn xx% | nnn xx% |
| All other causes | nnn xx% | nnn xx% |
| **Underlying causes of death** | N=XXX | N=XXX |
| Most common cause #1 | nnn xx% | nnn xx% |
| Most common cause #2 | nnn xx% | nnn xx% |
| Etc. |  |  |
| Most common cause #10 | nnn xx% | nnn xx% |
| All other causes | nnn xx% | nnn xx% |

**Table 14.** Protocol violations and deviations.

| Cause | Intervention<br>(N = ) | Usual Care<br>(N = ) |
| --- | --- | --- |
| **Any protocol violation or deviation** | nnn xx% | nnn xx% |
| **Any protocol violation** |  |  |
| **PV code and category** |  |  |
| Category 1 | nnn xx% | nnn xx% |
| Category 2 | nnn xx% | nnn xx% |
| Category 3... | nnn xx% | nnn xx% |
| **Any protocol deviation** |  |  |
| **PD code and category** |  |  |
| Category 1 | nnn xx% | nnn xx% |
| Category 2 | nnn xx% | nnn xx% |
| Category 3... | nnn xx% | nnn xx% |

**Table 15.**
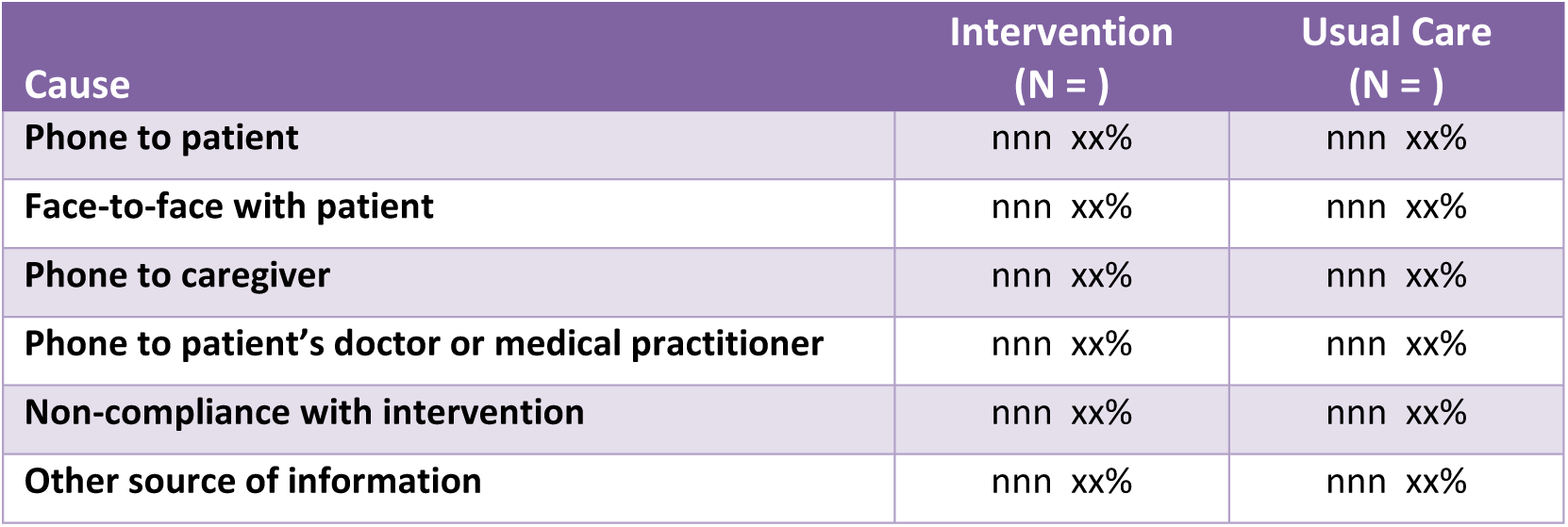
Form of assessment of 6-month outcomes.

### 6.2 Figures

**Figure 1.**
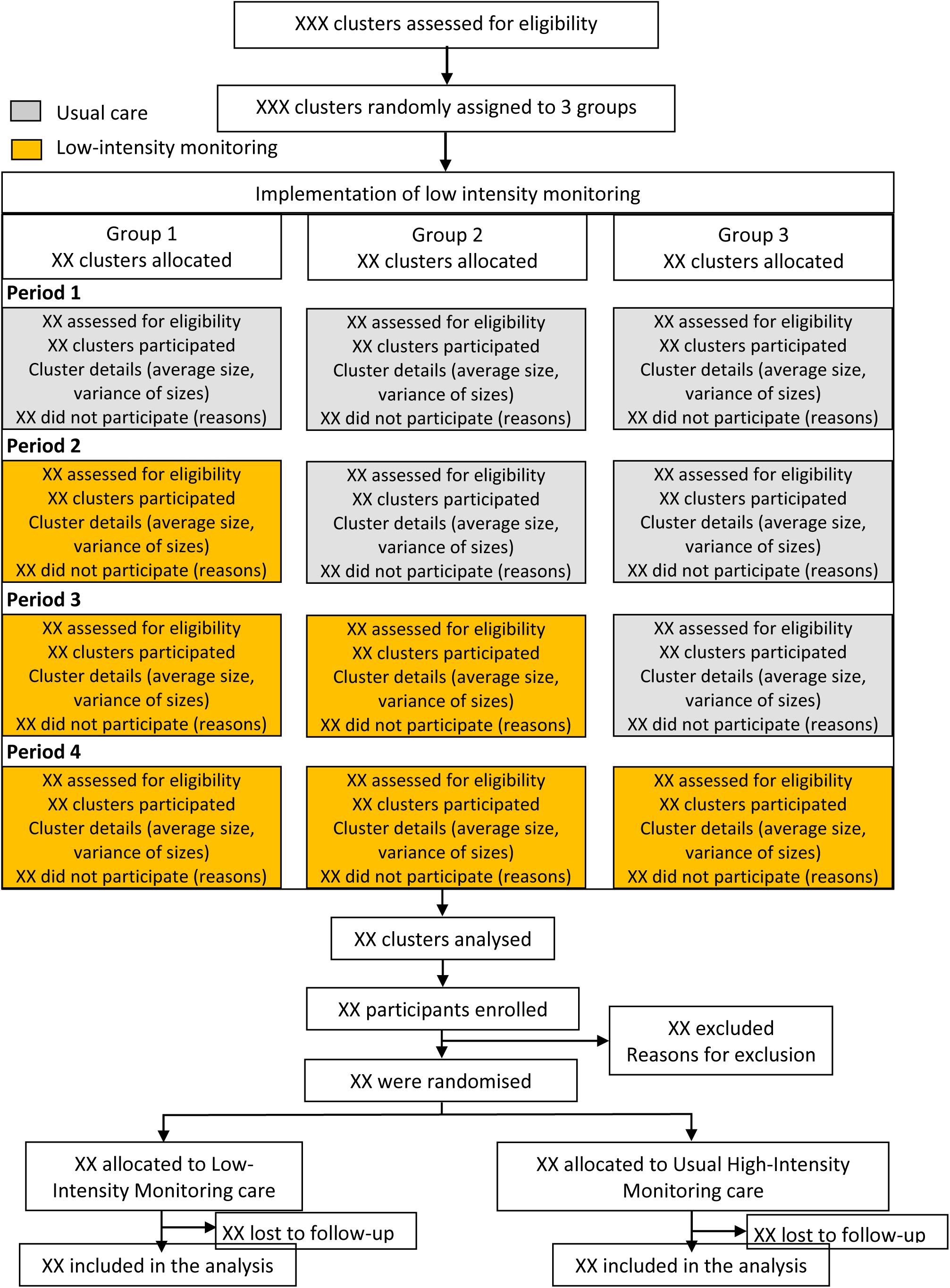
CONSORT diagram.

**Figure 2.**
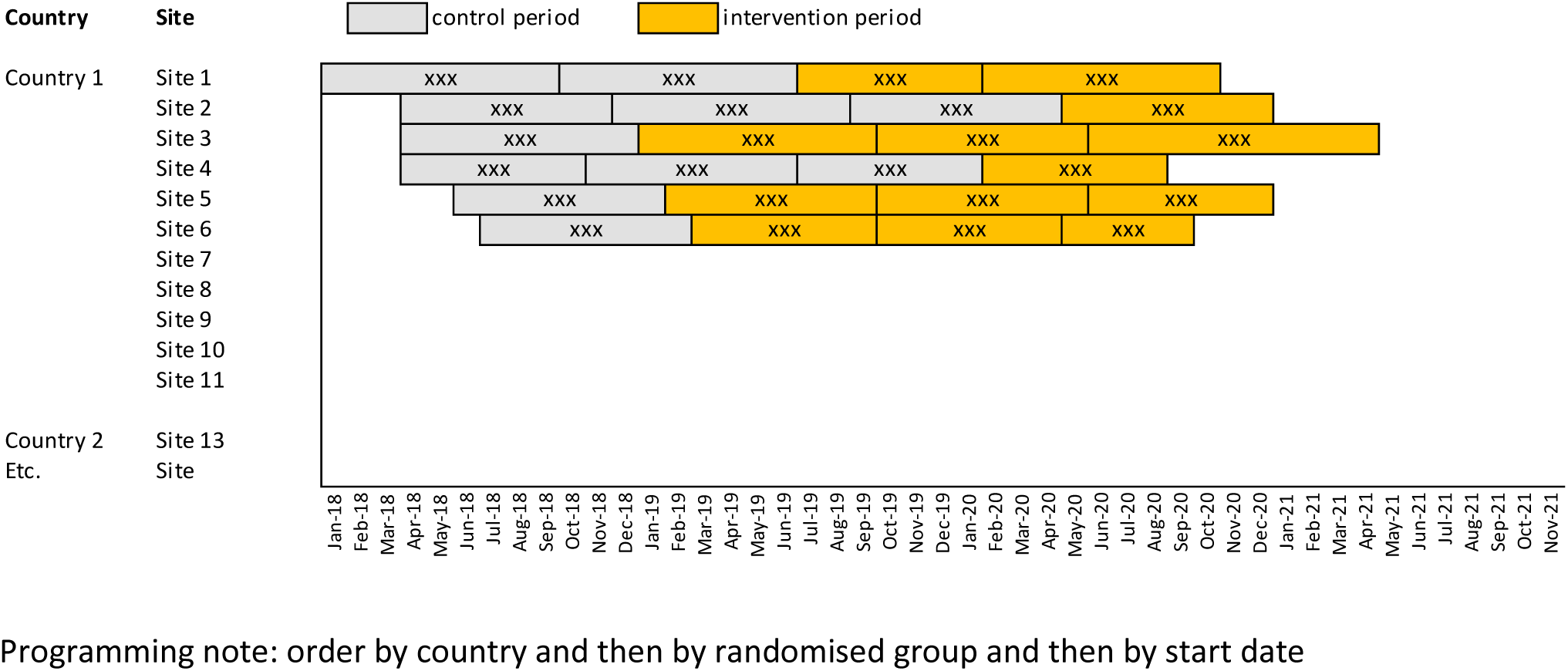
Start and stop dates of each period with number recruited per site per country

**Figure 3. Boxplot of systolic blood pressure over time**

Programming note: Display denominators (N) under the x-axis at key timepoints.

**Figure 4. Boxplot of diastolic blood pressure over time**

Programming note: Display denominators (N) under the x-axis at key timepoints.

**Figure 5. Boxplot of heart rate over time**

Programming note: Display denominators (N) under the x-axis at key timepoints.

**Figure 6. Grotta bar charts of mRS**

Programming note: Stacked bar chart with 2 bars (intervention vs control) per visit. Each bar to be of the same high (100%). Show the proportion in each category using labels on the bars.

**Figure 7: Boxplot of NIHSS by follow-up assessment**

**Figure 8. Forest plot for subgroup analysis of mRS at Day 90**

**Figure 9. Cumulative incidence function of time to hospital discharge over 7 days**

Programming note: add number at risk every day, median, quartiles, hazard ratio, 95% CI and P value from the Cox model.

**Figure 10. Kaplan Meier curve of mortality over 90 days**

Programming note: add number at risk every 10 days, median, quartiles, hazard ratio, 95% CI and P value from the Cox model.

**Figure 11. Bar chart of EQ-5D questions at 7 days**

**Figure 12. Bar chart of PREMs questions at 7 days**

**Figure 13. Bar chart of sleep-impairment questions at 7 days**

